# The Impact of Age on Gray Matter Volume Reduction in Anorexia Nervosa: A Systematic Review

**DOI:** 10.1101/2025.03.03.25322748

**Authors:** Huaze Gao, Shuo Chen, Lei Wang, Pei-an Betty Shih

**Affiliations:** Department of Psychiatry, University of California, San Diego, La Jolla, CA, United States; School of Engineering Science, Simon Fraser University, Burnaby, BC, Canada; Psychiatry and Behavioral Health, Neuroscience Ohio State University, Columbus, OH, United States

## Abstract

**Objective:** This study examines the relationship between gray matter (GM) volume reduction and age in individuals with Anorexia Nervosa (AN). Specifically, it investigates whether the magnitude and direction of GM volume differences between AN and healthy controls remain consistent across a range of age groups. Additionally, we reviewed regional GM alterations reported in the literature to characterize unique regional brain profiles observed in AN. By synthesizing neuroimaging studies and mean-age stratified analysis, this work provides insights into the possible impact aging can have on GM reduction in patients with AN.

**Methods:** Systematic review and meta-analysis were conducted using MRI-based neuroimaging studies assessing GM volume in AN patients and controls. A primary meta-analysis was run for all feasible studies combined, followed by a stratified analysis approach examining “younger mean-age” studies and “older mean-age” studies separately. Random effects models were used for the meta-analysis. Meta-regression was used to determine the influence of age on GM volume differences and was controlled for the body mass index to minimize the confounding effect recovery status has on the GM differences between groups. Regional GM alterations were reviewed and discussed.

**Results:** 44 studies, including 1391 individuals with AN and 1566 healthy controls, were included in the primary meta-analysis. No substantial heterogeneity was found across studies. Compared to their respective control groups, the younger-age studies, defined by studies with AN subject of mean age less than 18, exhibited greater significant GM volume loss (-5.39, 95% CI: -7.76 to -3.01, p<0.05) compared to older-age studies (-3.09, 95% CI: -4.16 to -2.03, p<0.05). Meta-regression subgroup results suggest that having older age in AN subjects is linked to less severe GM reduction relative controls. Our review of the regional GM literature reveals that alterations in the hippocampus, amygdala, and precuneus of the medial parietal lobe were more frequently reported than other brain regions in AN. In these regions, we also noticed that younger individuals with AN had more consistent volume reductions across studies, whereas studies with older AN showed greater variability.

**Conclusion:** Grey matter volume loss in AN is more pronounced in younger patients even after controlling for the effect of the recovery status. Having older age appears to contribute to less deficit in brain volume loss in AN, suggesting a protective mechanism underlying GM alteration in older AN patients. These findings reinforce the need for early intervention and prolonged recovery support and emphasize the need to develop lifespan-specific disorder management approaches. Future research should explore long-term GM recovery trajectories and the aging effect on GM alteration for older patients to refine strategies for neuroprotection in AN.

## I. Introduction

Anorexia Nervosa (AN) is a serious mental health condition marked by drastic food intake reduction, a profound fear of gaining weight, and a warped view of one’s body image. Initially described in the late 19th century as self-imposed starvation, AN remains one of the most life-threatening mental health conditions, exhibiting the highest mortality rates among psychiatric disorders. The disorder predominantly strikes adolescents and young adults, with peak onset occurring during adolescence. Although AN is significantly more prevalent in females, affecting approximately 0.3%-2% of women in the general population, it also occurs in males, albeit at substantially lower rates. Epidemiological studies indicate that up to 90% of diagnosed cases occur in females, with adolescent girls and young adult women constituting the most vulnerable demographic [1-3].

The etiology of AN is multifactorial, involving complex interactions between genetic, neurobiological, psychological, and sociocultural factors. While Western societies have historically reported higher prevalence rates due to pervasive thinness ideals, recent trends suggest a rising incidence of eating disorders in non-Western populations, likely driven by globalization and evolving beauty standards. Beyond its psychological and behavioral manifestations, AN has profound physiological consequences, affecting nearly every organ system. Among these, the central nervous system is particularly vulnerable, with prolonged malnutrition leading to significant structural and functional alterations in the brain [4, 5].

Advancements in neuroimaging, particularly magnetic resonance imaging (MRI), have facilitated a more comprehensive understanding of AN-related brain alterations. Studies show that AN with an active disorder (ill AN) is consistently associated with widespread reductions in brain volume (BV) with pronounced gray matter (GM) atrophy observed in cortical and subcortical regions [6, 7], while recovered AN (RecAN), defined most often by weight recovery, show evidence of partial GM volume recovery [18-21].

Some brain regions are more likely to be reported as altered in ill AN, such as the hippocampus, amygdala, parietal cortex, and anterior cingulate cortex [8-12]. These regions play key roles in memory processing, threat response, self-perception, and sensory integration, suggesting that their alterations may contribute to core features of AN, including heightened anxiety, distorted body image, and dysregulated appetite control [13-17]. However, most of the published studies utilized convenient clinical AN patients from treatment centers, predominantly adolescents and young adults, with a disproportionately low number of subjects in middle- and late adulthood. The narrow age range of AN in these studies implies that the results may not be generalizable to all AN patients in the community, ranging from adolescence to late life. The extent to which brain volume differences are differentially affected across different age groups remains unclear.

Of interest, neuroimaging studies have demonstrated that GM volume can partially recover following nutritional rehabilitation and weight recovery [18-21]. For example, short-term refeeding interventions led to measurable increases in brain volume, particularly within the first months of weight restorations [22, 23]. While these results suggest that the GM reduction improves with weight recovery in AN, full structural restitution remains inconsistent with some patients experiencing persistent deficits even after achieving weight normalization [24, 25]. Longitudinal studies indicate that brain volume recovery occurs most rapidly in the early stage of weight restoration but slows over time. Additionally, they found that older patients exhibit less cortical GM recovery in later treatment phases, suggesting that age has a notable influence on neuroplasticity [26].

Despite these insights, few studies have systematically compared GM volume alterations in AN across the different lifespans of AN patients. A previous meta-analysis that compared GM alterations between adolescent and adult AN did not directly probe the influence of age on these alterations [27]. Our study builds on this work using a larger dataset and applied complementary analyses to report the overall GM differences between AN and healthy controls and examine the effect age has on GM alteration by group stratification. Given that neurodevelopmental trajectories and brain plasticity likely differ across different lifespans, an improved understanding of the impact age has on GM volume alteration in AN is crucial for optimizing treatment strategies.

Prior research investigating subcortical regions, such as the hippocampus and amygdala in AN [8, 10, 28-31] did not provide adequate data to understand the influence of aging on regional alterations. This study undertook literature synthesis and meta-analyses to assess if AN-related GM global volume alterations may present differently in studies with younger-mean age AN compared to studies with older-mean age AN, thereby addressing a critical gap in knowledge regarding neurobiological effects of AN across the lifespan.

## II. Inclusion and Exclusion Criteria

All papers were retrieved prior to February 2025. PubMed, Google Scholar, Scopus, and Web of Science were used with appropriate keyword combinations. To ensure a comprehensive search, we used a combination of keywords related to Anorexia Nervosa (“anorexia nervosa” OR “eating disorder”) AND gray matter volume (“gray matter” OR “grey matter”) “brain volume” OR “MRI” OR “Magnetic Resonance Imaging” OR “regional”). We restricted our search for papers written in English only. A total of 421 papers were collected from the search, as shown in Figure 1. After removing duplicates, two reviewers, the authors of this study (HG, SC), independently screened the titles and abstracts to exclude studies that did not meet our inclusion criteria. Full-text articles were then assessed, and for the meta-analysis, studies were excluded if they did not provide numerical gray matter volume data suitable for inclusion. For the qualitative review of regional GM changes, studies were excluded if they did not investigate regional GM differences in individuals with AN compared to controls. Gray literature, case reports, reviews, editorials, and non-peer-reviewed studies were excluded from the study.

**Figure 1.**
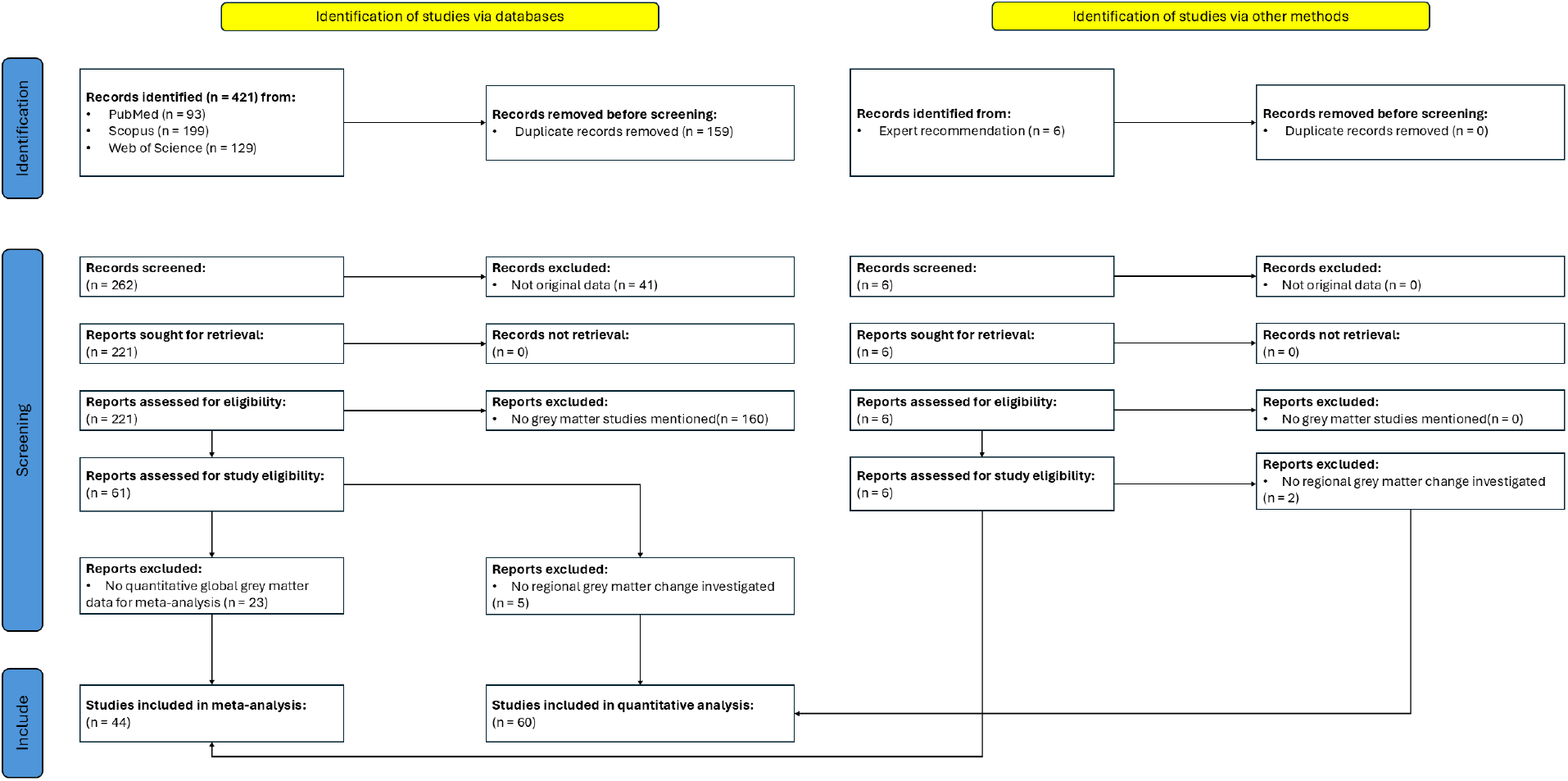
Flow diagram using Preferred Reporting Items for Systematic reviews and Meta-Analyses (PRISMA).

For meta-analysis, studies with missing age or gray matter volume data for either AN or healthy control groups were excluded. As shown in Table I, 44 papers remain after applying the exclusion criteria for meta-analysis. We used all eligible data from papers that included multiple case-control results with independent subjects. The papers used in the meta-analysis were sorted by the mean age of AN subjects. The number of AN patients and their age and body mass index (BMI) are reported.

**TABLE I:** Summary of GM Change (%) in AN meta-analysis study, sorted by mean age of AN subjects from youngest to oldest. # of AN = number of AN participants, Age of AN = mean age, BMI = body mass index, and GM Change (%) = grey matter change percentage with propagated standard deviation. All studies included for meta-analysis.

| DOI | Author/Year | # of AN | Age of AN | BMI of AN | GM Change (%) |
| --- | --- | --- | --- | --- | --- |
| 10.1371/journal.pone.0128548 | Fujisawa et al. 2015 [34] | 20 | 14.15 (1.81) | 14.35 (2.08) | -9.98 (-1.07) |
| 10.1016/j.jpsychires.2008.03.013 | Castro-Fornieles et al. 2009 [9] | 12 | 14.50 (1.50) | - | -14.59 (-1.52) |
| 10.2147/NDT.S73239 | Bomba et al. 2015 [22] | 11 | 14.63 (2.77) | 18.87 (1.86) | -9.15 (-0.12) |
| 10.1002/eat.22805 | Olivo et al. 2018 [35] | 22 | 14.90 (1.60) | 19.30 (2.00) | -0.97 (-2.22) |
| 10.1192/j.eurpsy.2020.19 | Curzio et al. 2020 [36] | 24 | 15.17 (2.00) | 14.50 (1.70) | -4.41 (-0.94) |
| 10.1016/j.pscychresns.2010.06.007 | Gaudio et al. 2011 [37] | 16 | 15.20 (1.70) | 14.20 (1.40) | -7.44 (-0.09) |
| 10.1016/s0022-3476(96)70021-5 | Katzman et al. 1996 [38] | 13 | 15.40 (1.20) | 15.60 (1.70) | -1.38 (-0.15) |
| 10.1016/j.jaac.2013.07.007 | Frank et al. 2013-1 [39] | 19 | 15.40 (1.40) | 16.20 (1.10) | -0.71 (-0.09) |
| 10.1016/j.jpsychires.2015.06.019 | Seitz et al. 2015 [5] | 56 | 15.47 (1.43) | 15.13 (1.37) | -7.08 (-0.88) |
| 10.1097/PSY.0b013e31824ef10e | Mainz et al. 2012 [40] | 19 | 15.70 (1.50) | 15.30 (1.50) | -18.02 (-3.59) |
| 10.1016/j.pscychresns.2017.06.011 | Gaudio et al. 2017 [41] | 14 | 15.70 (1.60) | 16.20 (1.20) | -4.75 (-0.43) |
| 10.1016/j.pscychresns.2018.10.007 | Myrvang et al. 2018 [30] | 30 | 15.80 (1.70) | 16.30 (1.60) | -7.68 (-0.91) |
| 10.1002/eat.22161 | Lazaro et al. 2013 [20] | 35 | 16.30 (1.30) | 19.30 (1.10) | -0.22 (-0.02) |
| 10.1016/j.pnpbp.2025.111248 | Wronski et al. 2025 [42] | 131 | 16.41 (3.03) | 14.64 (1.42) | -6.13 (-0.65) |
| 10.1111/pcn.13626 | Bahnsen et al. 2024 [8] | 165 | 16.44 (3.06) | 14.69 (1.39) | -5.94 (-0.66) |
| 10.1017/S0033291724000394 | Keeler et al. 2024 [43] | 82 | 16.83 (3.22) | 15.03 (1.23) | -3.00 (-0.33) |
| 10.1001/archpedi.1997.02170450043006 | Katzman et al. 1997 [24] | 6 | 17.00 (1.40) | 23.00 (3.70) | 3.96 (0.45) |
| 10.1001/archpsyc.1997.01830180055006 | Lambe et al. 1997 [25] | 12 | 18.90 (6.90) | 20.50 (2.30) | 1.60 (0.22) |
| 10.1016/j.pscychresns.2018.08.009 | Yue et al. 2018 [44] | 35 | 19.34 (4.09) | 15.34 (1.96) | -6.03 (-0.59) |
| 10.1016/j.pscychresns.2013.03.010 | Amianto et al. 2013 [45] | 17 | 20.00 (4.00) | 16.00 (1.00) | -0.18 (-0.02) |
| 10.1542/peds.2008-0170 | Chui et al. 2008 [18] | 66 | 21.30 (2.30) | 21.80 (3.40) | 0.32 (0.03) |
| 10.1016/j.pscychresns.2010.12.008 | Boghi et al. 2011A [46] | 10 | 21.40 (2.50) | 14.95 (1.16) | -2.60 (-0.25) |
| 10.1002/eat.22397 | Burkert et al. 2015 [28] | 21 | 21.60 (5.70) | 15.80 (1.30) | -8.80 (-1.11) |
| 10.1038/s41398-020-0809-7 | Kaufmann et al. 2020 [26] | 24 | 22.25 (4.08) | 14.25 (1.08) | -5.14 (-0.51) |
| 10.1176/appi.ajp.2007.06111861 | Muhlau et al. 2007 [47] | 25 | 22.30 | 24.80 | -2.51 |
| 10.1503/jpn.140193 | Bar et al. 2015 [48] | 26 | 22.96 (4.97) | 16.97 (1.46) | -7.76 (-0.97) |
| 10.1017/S0033291713002389 | Fonville et al. 2014 [49] | 31 | 23.00 (10.00) | 15.80 (1.40) | -3.50 (-0.26) |
| 10.1176/appi.ajp.2013.12101294 | Frank et al. 2013-2A [19] | 19 | 23.10 (5.80) | 16.00 (1.10) | 3.51 (0.40) |
| 10.1016/j.biopsych.2005.06.014 | Wagner et al. 2006 [21] | 14 | 23.70 (5.30) | 21.20 (2.00) | 2.82 (0.27) |
| 10.1002/eat.22918 | Nickel et al. 2018A [50] | 34 | 23.80 (4.30) | 16.10 (1.40) | -5.97 (-0.70) |
| 10.1016/j.neuroimage.2011.09.042 | Friedrich et al. 2012A [51] | 12 | 24.30 (6.20) | 15.90 (1.60) | -7.87 (-1.01) |
| 10.1016/j.neuroimage.2011.09.042 | Friedrich et al. 2012B [51] | 13 | 25.00 (4.80) | 19.50 (1.40) | -3.67 (-0.50) |
| 10.1016/j.pscychresns.2010.02.004 | Joos et al. 2010 [52] | 29 | 25.00 (4.80) | 16.00 (1.20) | -4.02 (-0.20) |
| 10.1186/s40337-021-00466-w | Brodrick et al. 2021A [53] | 37 | 25.40 (6.75) | 17.83 (1.17) | -3.51 |
| 10.1002/eat.20549 | McCormick et al. 2008 [54] | 18 | 25.60 (7.24) | 13.50 (2.14) | -4.51 (-0.51) |
| 10.1016/j.pscychresns.2020.111138 | Oliva et al. 2020 [55] | 15 | 25.87 (6.15) | 20.10 (2.04) | -1.15 (-0.13) |
| 10.1186/1471-244X-11-179 | Brooks et al. 2011 [56] | 14 | 26.00 (1.90) | 15.60 (0.40) | -3.97 (-0.15) |
| 10.1002/eat.10111 | Swayze et al. 2003 [57] | 18 | 26.00 (7.60) | 24.20 (3.30) | -2.46 (-0.31) |
| 10.1016/j.bbr.2009.08.035 | Suchan et al. 2010 [58] | 15 | 26.80 (8.40) | 16.00 (1.30) | -2.54 (-0.40) |
| 10.1002/eat.20840 | Roberto et al. 2011 [59] | 32 | 26.91 (6.41) | 20.01 (0.59) | -2.68 (-0.35) |
| 10.3389/fpsy.2017.00030 | Scaife et al. 2017B [12] | 14 | 27.00 (6.50) | 20.90 (1.60) | -1.86 (-0.09) |
| 10.1002/eat.22918 | Nickel et al. 2018B [50] | 24 | 27.10 (7.00) | 20.60 (1.30) | -3.23 (-0.35) |
| 10.1503/jpn.130135 | Via et al. 2014 [60] | 19 | 28.37 (9.55) | 17.03 (1.09) | -1.37 (-0.14) |
| 10.1016/j.pscychresns.2017.07.004 | Kohmura et al. 2017 [61] | 23 | 28.50 (6.70) | 13.20 (1.50) | -9.74 (-0.99) |
| 10.1186/s40337-021-00466-w | Brodrick et al. 2021B [53] | 32 | 29.40 (8.27) | 22.09 (2.33) | -1.24 |
| 10.3389/fpsy.2017.00030 | Scaife et al. 2017A [12] | 12 | 29.40 (6.00) | 15.40 (1.90) | -6.98 (-0.33) |
| 10.1002/erv.1060 | Joos et al. 2011 [62] | 5 | 29.60 (5.10) | 19.90 (1.50) | -1.61 (-0.07) |
| 10.1176/appi.ajp.2013.12101294 | Frank et al. 2013-2B [19] | 24 | 30.30 (8.10) | 20.80 (2.40) | 1.54 (0.19) |
| 10.1016/j.pscychresns.2010.12.008 | Boghi et al. 2011B [46] | 11 | 35.91 (9.25) | 16.01 (2.07) | -3.59 (-0.39) |
| 10.1016/J.PSCYCHRESNS.2021.111393 | Mishima et al. 2021 [63] | 35 | 36.30 (10.00) | 14.20 (2.50) | -5.81 (-0.60) |

For the qualitative review, we focused on regional GM volume differences in AN compared to healthy controls. Studies that exclusively investigated cortical thickness, surface area, or white matter were excluded. We identified and reviewed all studies that examined regional GM alterations in AN, their associations with AN-related clinical measures, and discussed their potential contributions to the underlying mechanisms of AN.

## II. Methods

The GM volume change percentage is calculated based on the reported GM volume for AN and HC. Given the GM mean of AN and HC to be µ_AN_ and µ_HC_, and the standard deviation of AN and HC to be σ_AN_ and σ_HC_, we obtain the mean and standard deviation of GM change in percentage µ_GM_ and σ_GM_ as:

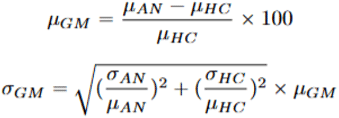

Meta-analyses were conducted using the *metafor* package in R, employing a random-effects model to generate forest plots. For studies without mean and standard deviation values, imputation from the group average will be applied. To evaluate the correlation between GM change and age progression within each age group, meta-regression analysis was performed using semi-partial regression implemented in the *pingouin* package [33] in Python. Mean-BMI was included as a covariate in the regressions to account for the effect AN recovery has on GM volume.

Additional review was conducted for GM regional differences. After screening, 67 studies were identified to contain data for regional GM changes in individuals with AN compared to healthy controls. Among these, 50 studies investigated global GM changes, with 44 (88%) used in the meta-analysis. Of these 50 studies, 43 further reported data on regional GM differences between AN and controls. 37(86.05%) of these studies were used in the global GM meta-analysis. Additional 17 studies focused solely on regional GM changes without assessing global GM were utilized to extract data for regional differences. The qualitative review of regional GM changes is based on 60 studies that examined regional GM differences between AN and controls.

## IV. Results and Discussion

### 1) Total GM Volume Meta-Analysis

Forty four (44) studies with a total of 1391 AN patients and 1566 healthy controls were included in our meta-analysis. These studies were stratified by AN’s mean-age to two sub-groups. Seventeen (17) studies had a mean-age in AN of less than 18 years (i.e., younger age group), while 27 studies had mean-age in AN of equal to or greater than 18 years (i.e., older age group). Figure 2 presents the distribution of GM change in both groups, illustrating distinct GM volume reduction across both age groups. The summary effect size estimates indicate that AN in the younger group studies (<18 years old) exhibit a more significant overall GM volume reduction, with a mean change of -5.39 (95% CI: -7.76 to -3.01), compared to -3.09 (95% CI: - 4.16 to -2.03) in the older age group (≥18 years old). Both age groups demonstrate statistically significant GM atrophy, with p-values below 0.05, underscoring the robustness of these findings.

**Fig. 2:**
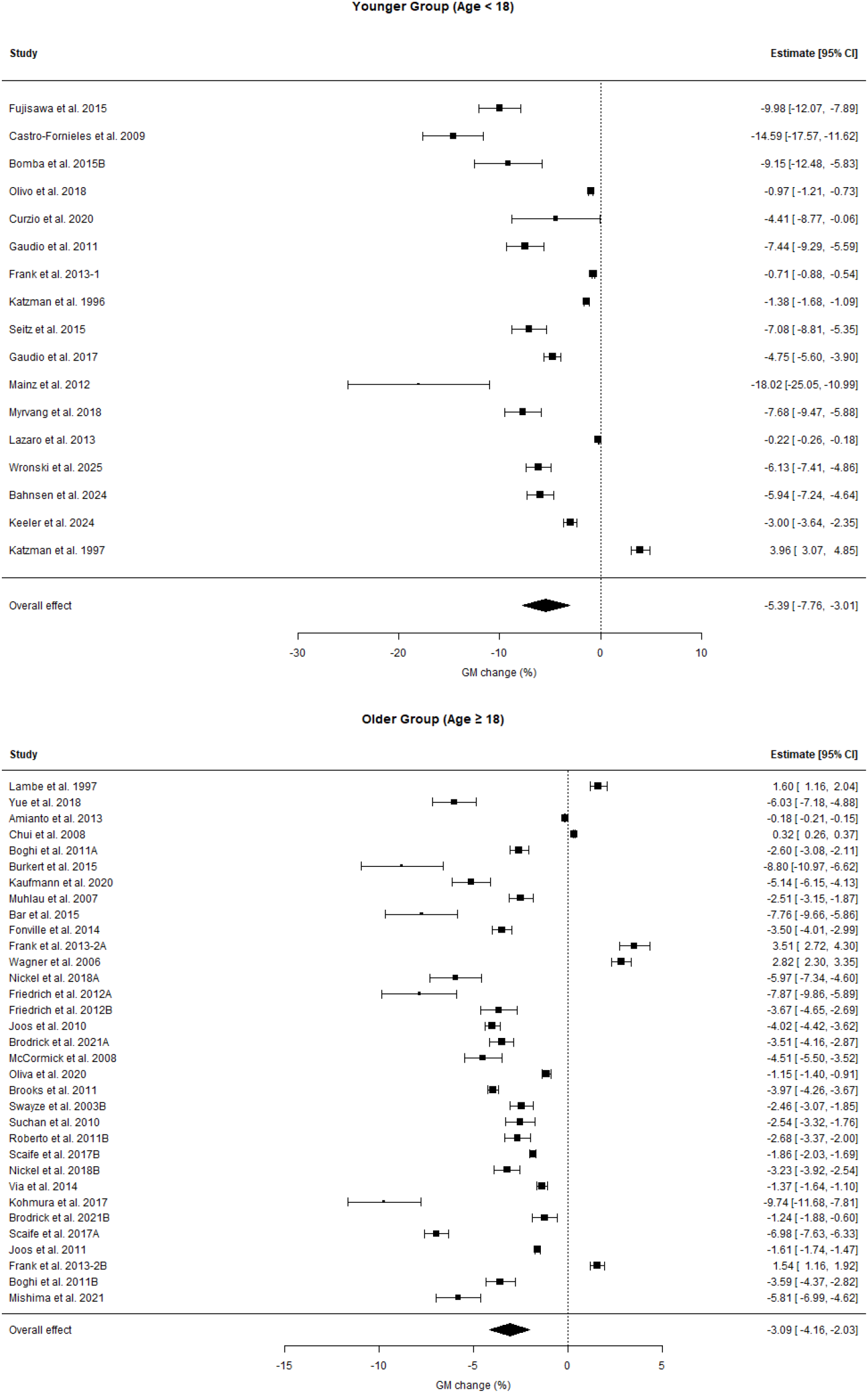
Forest plot depicting GM change (%) in younger and older groups with a cut-off age of 18. Studies are sorted top down by ascending mean age within each group. Each horizontal line represents the GM change with its standard deviation, while the filled cube marker indicates the mean GM change for each study. The studies with missing standard deviations of GM change are imputed by the group mean. The overall effect size is shown as a diamond shape at the bottom of each plot. The dashed vertical line at zero serves as a reference, indicating no change. The estimated 95% confidence interval is listed on the right side. Letters after the study year indicate that the study included multiple distinct groups of individuals with AN.

Heterogeneity analysis using Cochran’s Q indicated no significant heterogeneity in either the younger group (Q = 4.0191, p = 0.9995) or the older group (Q = 3.3845, p = 1.0000).

A meta-regression analysis was conducted to explore the influence of age on GM volume reduction (AN versus controls). Figure 3 demonstrates the correlational relationships between age and GM change within each mean-age-stratified group. We found a more significant correlation between age and GM volume reduction in AN in the younger age studies compared to the older age studies. In essence, the older the AN’s age, the less GM reduction there is. We illustrate this pattern using two cut-off age thresholds of 18 (Figure 3, Top) and 26 (Figure 3, Bottom). Age less than 18 represents adolescence in lifespan, whereas 26 was selected using the 75th percentile age point of all studies. In the younger groups, the regression coefficients are 0.54 (p = 0.026) and 0.27 (p = 0.116) for the cut-off ages of 18 and 26, respectively. In the older groups, regression coefficients are -0.05 (p > 0.05) and 0.2 (p > 0.05) for the cut-off ages of 18 and 26, respectively. The comparisons between younger and older groups in the two age cut-off analyses show that older age had the most significant protective effect on GM reduction for AN in studies with mean age of less than 18. Regression correlations in the older group analyses suggest that age’s protective effect on GM reduction may persist despite having inadequate power to show statistical significance.

**Fig. 3:**
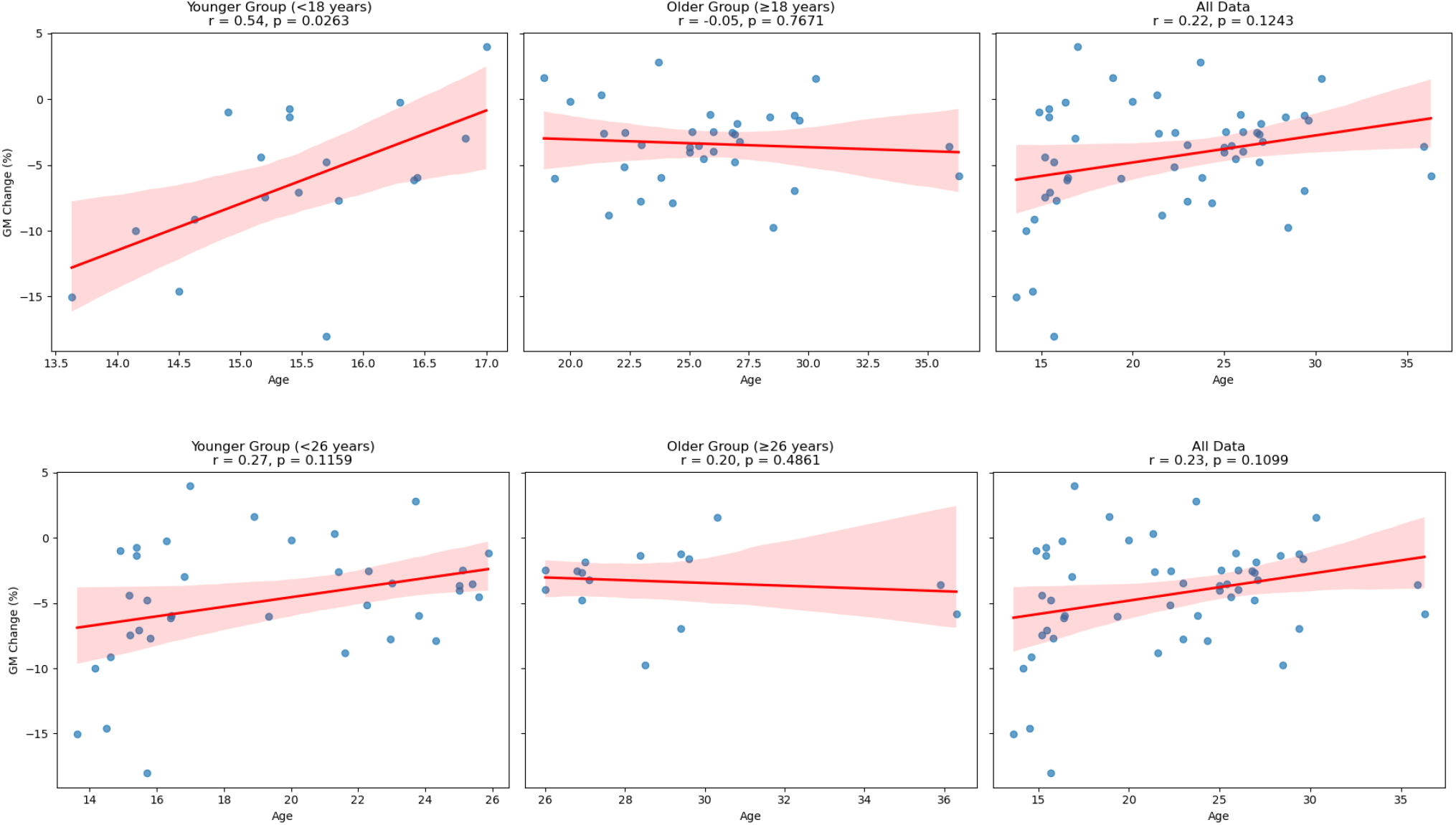
Meta-regression plots illustrating the relationship between Age (Mean) of AN subjects and GM Change (%) compared to control group, adjusted for AN’s BMI (Mean). Analysis was stratified by mean-age of AN in studies: younger group (Left), older group (Middle), and all studies combined (Right). The top row represents the results with a cut-off age of 18, and the bottom row shows the results with a cut-off age of 26. Scatter points represent individual study data, while the red lines depict the meta-regression fit. The shaded regions indicate the 95% confidence intervals.

### 2) Regional GM

We identified three brain regions that were most frequently reported to show GM alterations in individuals with AN: the hippocampus, amygdala, and precuneus. Below, we summarize the findings for each region, considering pattern differences between studies with younger (<18 years) and older (≥18 years) AN subjects (see Table II for a detailed summary of study findings).

**TABLE II:**
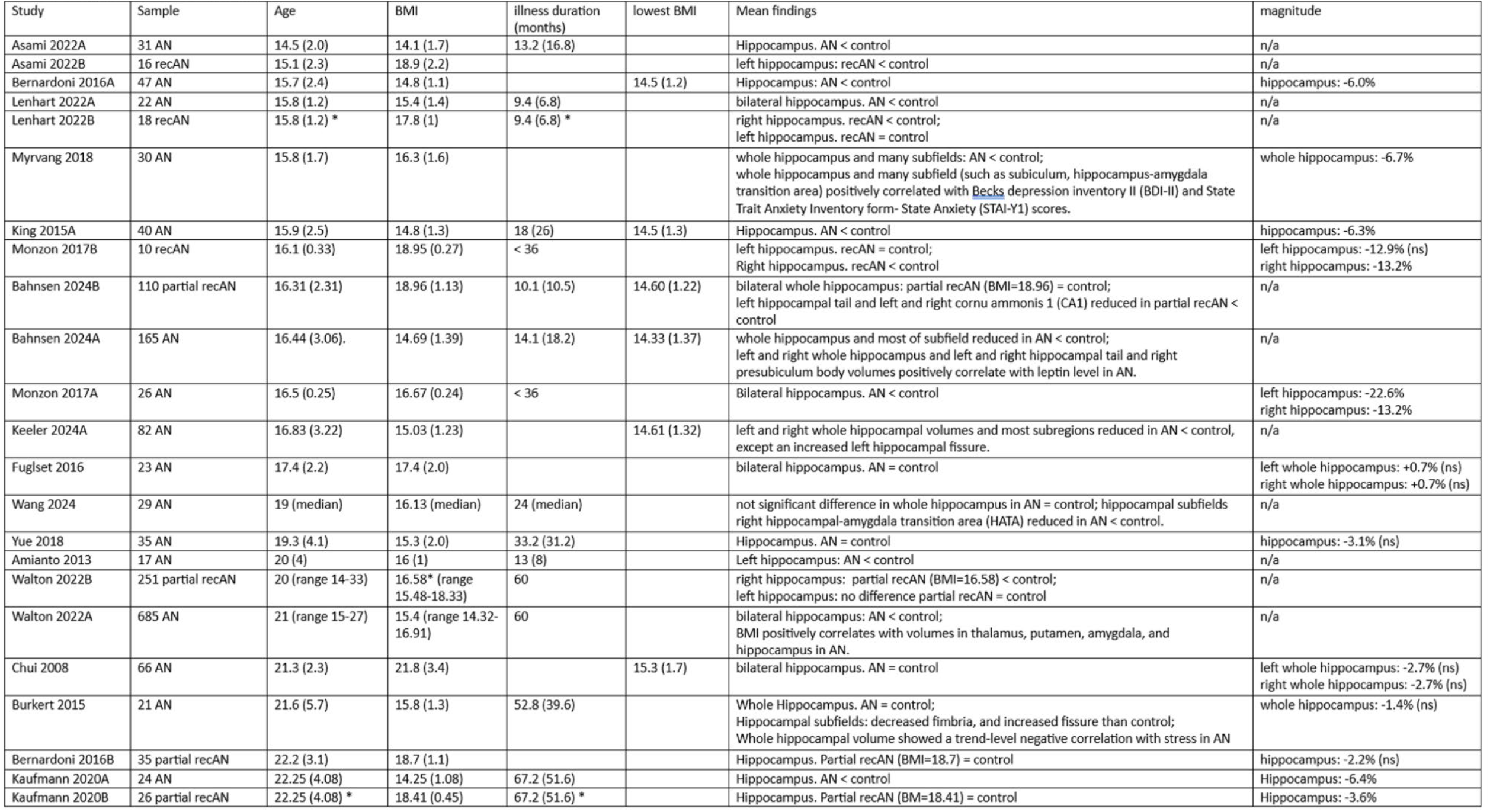
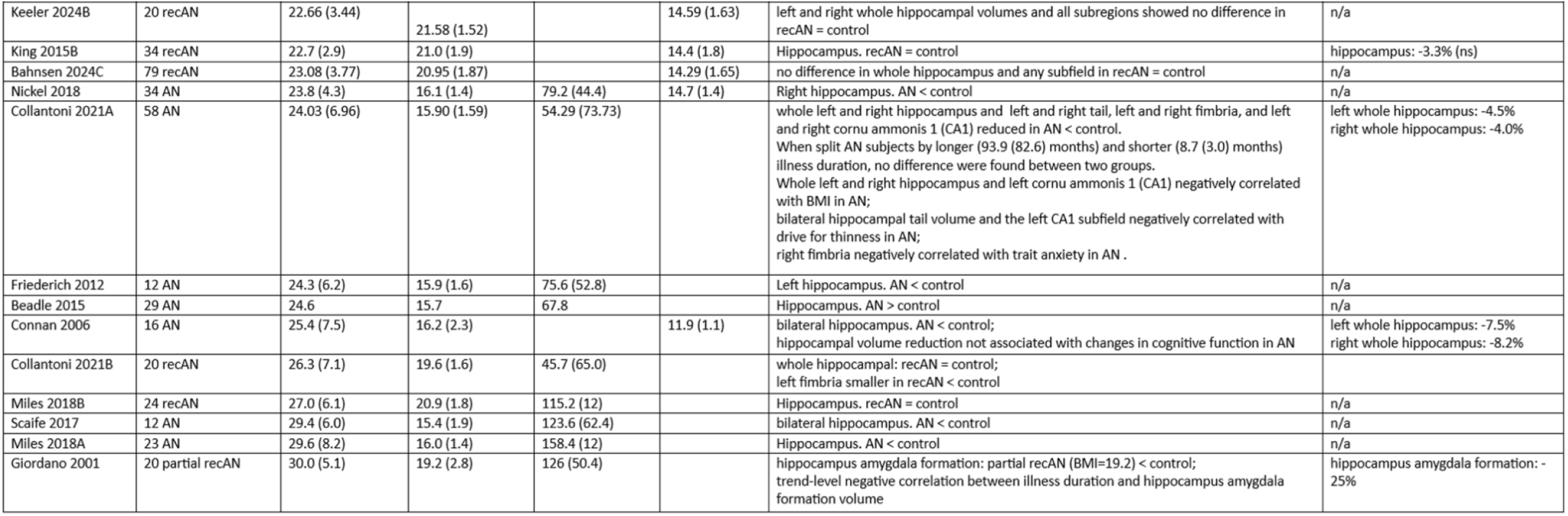
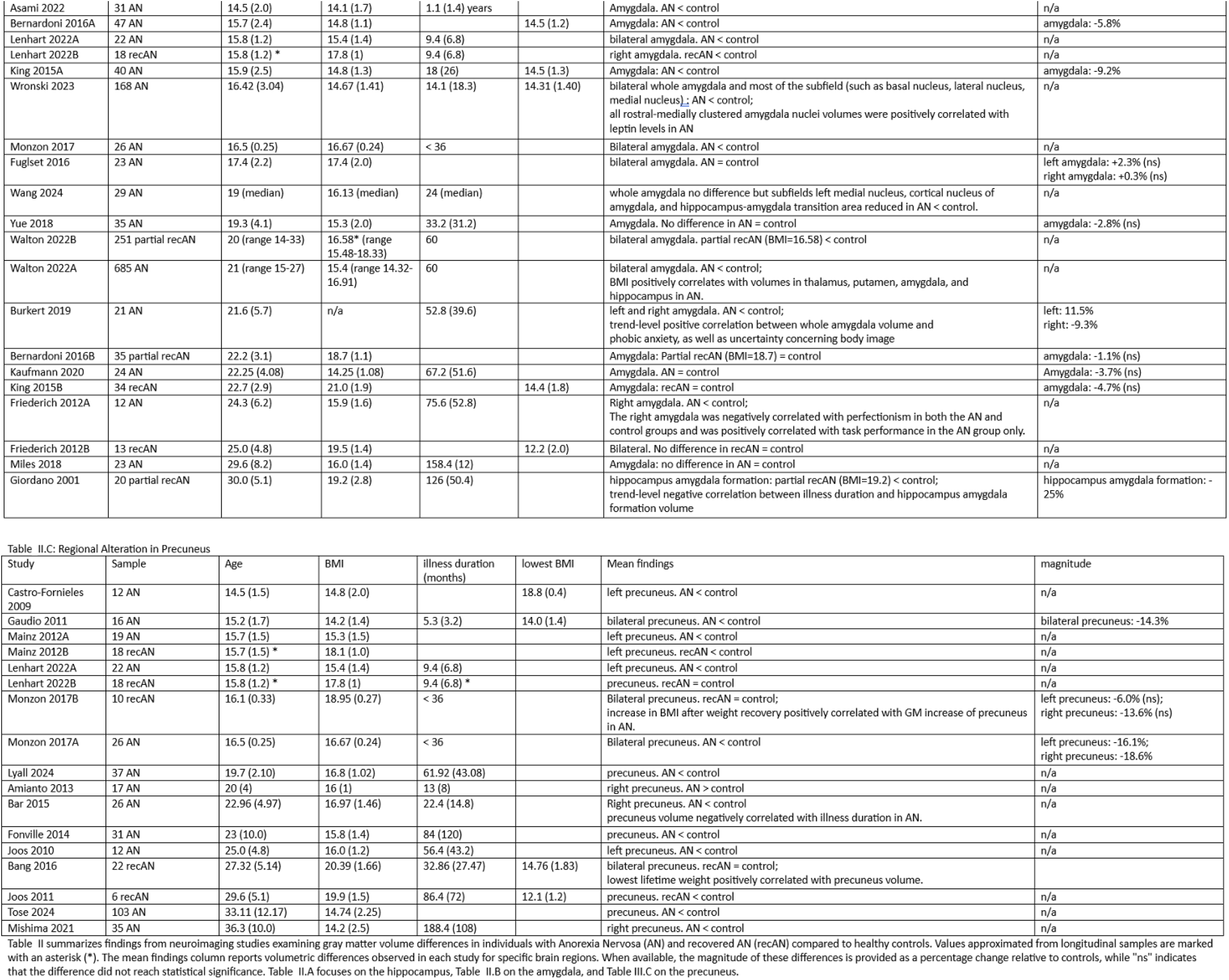
A:Regional Alteration in Hippocampus.

Among the 60 studies reviewed, 24 compared hippocampus volume in acute AN to controls. Studies conducted in younger AN showed that eight out of nine studies reported reduced hippocampal volume in individuals with acute AN compared to healthy controls[4, 8, 11, 30, 64-66]. One study [67] found no significant difference; however, the author of this study mentioned that they included 11 out of 23 AN participant who were partially weight-recovered (BMI>17.5), which may have contributed to the null findings. In studies conducted with older AN, 10 out of 15 studies reported reduced hippocampal volume in AN [10, 12, 23, 26, 29, 45, 50, 51, 68, 69]. Four studies [18, 28, 31, 44] found no significant difference, while one study [70] reported increased hippocampal volume in AN.

Fifteen (15) studies compared amygdala between acute AN and control. Findings in studies with younger AN largely mirrored those for the hippocampus. Six out of seven studies reported reduced amygdala volume in acute AN [4, 11, 64-66, 71]. The remaining study [67] found no significant difference, which, as noted earlier, included partially weight-recovered AN participants, potentially influencing the overall result. In the studies with older acute AN, findings were more variable. Four studies reported reduced amygdala volume in AN [23, 29, 51, 72], while the remaining four studies found no significant difference [26, 31, 44, 69].

Findings in the precuneus were largely consistent across studies. Among the 13 studies, 12 reported reduction in acute AN compared to controls. In studies with younger participants, all five studies reported reduced precuneus volume in acute AN [9, 11, 37, 40, 66]. Similarly, in the older AN, six out of seven studies also found reduced precuneus volume [48, 49, 52, 63, 73, 74]. However, one study reported increased precuneus volume in AN compared to control [45].

These regions were not only the most frequently reported regions with altered GM volume in AN, but also exhibited a pattern where younger individuals with acute AN were more consistently found to have reduced GM volume. In comparison, findings in studies with older AN show greater variability. Older age appears to normalize or protect against regional GM atrophy in AN.

## V. Discussion

To our knowledge, this is the largest meta-analysis to date examining GM volume changes in AN, and the first one to have stratified analysis by the mean age of AN subjects. Our findings provide robust evidence of significant GM volume reduction in individuals with AN, observed in both younger and older age groups. The age-stratified results show that the magnitude of GM reduction is greater in younger individuals with AN, or less severe in AN of older age. Controlling for the confounding effect of AN recovery, our regression analysis further shows a significant protective effect older age has on the GM volume reduction, especially in studies using AN with a mean age of less than 18. This finding reinforces the idea that age plays a critical role in the extent of brain volume changes associated with the disorder. This finding aligns with a previous meta-analysis [27], which reported significantly greater GM reduction in adolescents compared to adults with acute AN. Despite this adjustment, the pattern of more pronounced GM reduction in younger AN remained evident, suggesting that age-related differences in GM atrophy are not solely attributable to acute undernutrition but may also reflect underlying neurodevelopmental vulnerability. Furthermore, our meta-analysis, which included a larger sample size, strengthens the reliability of these findings. Additionally, the absence of significant heterogeneity across studies suggests a degree of consistency in reported GM alterations, further supporting the robustness of our results.

Adolescence represents a critical period of neurodevelopment, during which individuals are particularly vulnerable to GM volume and structural changes. Both risk factors, such as malnutrition and trauma, and protective factors, including balanced nutrition and a healthy lifestyle, can have a greater impact on the brain [75]. The protective effect older age has on GM reduction is preserved in regression slopes in the older AN group analysis, but was not statistically significant. This suggests that GM volume may be more stable and resilient to damage by risk factors in adulthood. Dietary and behavior factors contribute significantly to health outcomes as we age. Identifying the protective dietary nutrition and behaviors for the brain is particularly relevant in the prevention of relapse in AN.

The current literature on GM alteration in AN reports data of adolescent and young-adult patients. Expanding research criteria to intentionally include older individuals with AN, particularly those over the age of 40, is critical. Only by including patients across all lifespans can we achieve an understanding of GM volume trajectories in all stages of life and discover protective factors in patients with AN. A balanced representation of AN from all age groups will also yield results that are more generalizable.

Based on our review, volume alterations for specific regions have been reported more frequently in AN than others. The hippocampus and amygdala, two core limbic structures, have attracted significant interest and are frequently reported to exhibit volumetric alterations in AN. The hippocampus is well known for its crucial role in memory, learning, and emotion processing [13, 14, 16]. This essential brain region had been found to have a reduced volume in AN by many studies [4, 8, 10, 11, 23, 26, 28-30, 43, 45, 50, 51, 64-66, 68, 69]. However, some studies reported no significant differences [31, 44] and increased volumes compared to controls [70].

Regarding age-related differences, studies with younger AN subjects appear more likely to report more significant hippocampal volume reduction compared to studies with older subjects (Myvang 2018 with age 15.8 has -6.7%; King 2015 with age 15.9 has -6.3%; Collantoni with age 21.6 has left: -4.5%, right: -4.0%.) [7, 10, 11, 28, 30, 65]. This pattern is consistent with our meta-analysis findings on global GM volume alterations. It suggests that younger individuals with AN may experience more severe regional brain atrophy.

A study conducted with adolescent subjects found that the volume of the whole hippocampus and several hippocampal subfields in AN positively correlated with depression and anxiety symptoms, including the hippocampus-amygdala transition area and Beck’s Depression Inventory-II (BDI-II) [11]. However, two other studies observed the opposite. One found a significant negative correlation between bilateral hippocampal tail volume and the left cornu ammonis 1 (CA1) subfield with drive for thinness, as well as between the right fimbria and trait anxiety [10]. The other study reported a trend-level negative association between total hippocampal volume and stress levels, though it did not find significant hippocampal volume differences between AN patients and controls, except for a smaller fimbria and a larger hippocampal fissure in AN patient [28]. One study of hippocampal volume about illness duration reported a trending negative correlation between illness duration and the hippocampus-amygdala formation volume in the AN group [29]. However, more recent studies attempting to replicate this finding have not observed significant associations between illness duration and neither hippocampal or amygdala volume [10, 71, 76].

The extent to which hippocampal volume normalizes with weight recovery remains inconsistent in the literature. Some studies have reported persistent hippocampal volume reductions after weight recovery [11, 64, 66], whereas a more significant number of studies have found evidence of normalization [4, 8, 26, 43, 65, 69]. One of the most recent longitudinal studies with a large sample size (AN = 165) explicitly examined hippocampal subregions [8]. It found that while the whole hippocampus and most subfields were reduced in acute AN, patients who achieved partial weight recovery (BMI = 18.96) at follow-up exhibited a significant increase in whole hippocampal volume compared to when they were acutely ill. Furthermore, at the second time point, there were no significant differences in whole hippocampal volume or most hippocampal subfields (including the hippocampus-amygdala transition area) between partially recovered AN patients and controls [8]. Another longitudinal study further supported these findings, demonstrating that the amygdala and hippocampus exhibited volume reductions during the underweight phase, but these volumes normalized following weight recovery [26].

Amygdala is a brain region that has been primarily reported to be reduced in volume in acute AN across multiple studies [4, 11, 23, 29, 51, 64-66, 71, 72, 77]. However, a few studies have reported no significant differences in amygdala volume in AN compared to controls [31, 44, 69].

Amygdala is known for its role in the fear and reward systems [17, 78] and contributes to both aversive and appetitive responses [79, 80], which are thought to play a critical role in AN through both physiological and psychological pathways. The amygdala transmits information to various brain structures, including the hippocampus, and its interactions with the hippocampus may support the conversion of sensory inputs into behavioral responses [81].

Notably, in a study that found no reduction in total amygdala volume in AN, atrophy was still observed in specific subfields, such as the right hippocampal-amygdala transition area, suggesting volume loss in at least some parts of the amygdala [31]. Similarly, another study reported no difference between acute AN and HC. At the same time, an increased amygdala volume was observed in cross-sectionally recovered AN compared to the acute AN group, suggesting that amygdala abnormalities in AN may be influenced by underweight conditions [69]. Moreover, a recent large-scale study (N = 685 AN) reported a positive correlation between BMI and amygdala and hippocampal volumes, further supporting the idea that malnutrition and underweight conditions may contribute to amygdala atrophy in AN [23].

The relationship between amygdala volume and clinically relevant AN measure has also been explored. One study measuring hormone levels in AN found that both amygdala and hippocampal volumes were significantly positively correlated with follicle-stimulating hormone (FSH) levels, suggesting a possible relationship between these regional volumes and sex hormone recovery [40]. Additionally, leptin levels were found to be associated with amygdala nuclei in the rostral-medial cluster, which the authors suggested may indicate that olfactory and food-related reward subregions are linked to hypoleptinemia in AN [42]. Further, one study reported a negative correlation between right amygdala volume and perfectionism scores [51], while another study observed a trend-level positive correlation between whole amygdala volume and phobic anxiety, as well as uncertainty concerning body image [72].

Functional imaging studies have found that, compared to controls, AN patients exhibit hyperactivation in the amygdala when viewing food images [82] and when hearing negative words related to body image [83]. Furthermore, a recent meta-analysis reported increased resting-state activity in the right parahippocampal gyrus and amygdala in AN [84]. These amygdala structural and functional abnormalities suggest that individuals with AN may perceive food- and body-related cues as fear-inducing stimuli. The persistent hyperactivity of this brain region may further contribute to the drive for thinness trait and restrictive eating behaviors.

The literature suggests that amygdala atrophy tends to normalize with weight restoration [4, 51, 65]. Although some studies have reported persistent amygdala reduction, many appear to have focused on partially weight-recovered subjects [29, 66]. For instance, one of the earliest studies found a significant 25% reduction in hippocampus-amygdala formation volumes in recovering AN patient with a mean BMI of 19.4 [29]. Similarly, another longitudinal study reported persistent proper amygdala reduction at the second scan in AN patient with a low mean BMI of 17.8 [66]. Therefore, this region’s structural abnormalities seem to be recoverable with weight gain.

Precuneus is another brain region that has been frequently reported as altered in AN. Located in the medial parietal lobe, precuneus is involved in voluntary attention shifts, visuospatial imagery, and personal identity processing [13, 15]. Similar to other regions previously discussed, the precuneus is reduced in AN across multiple studies [9, 11, 37, 40, 45, 48, 62, 66, 74, 77, 84, 85], with one exception reported by Amianto et al. [45], which found an increased volume in AN.

One study identified a negative correlation between AN illness duration and GM volume in precuneus [48] regarding the association between the precuneus and clinical measures. Another study found that the lowest lifetime weight was positively correlated with precuneus and insula volume in long-term recovered AN individuals (lowest BMI: 14.76), despite the absence of significant differences in total and regional GM volume compared to HC [76]. These findings seem consistent with a previous study conducted on long-term recovered AN individual (average illness duration: 7.2 years, lowest BMI: 12.1), which demonstrated persistently reduced precuneus volume compared to HC even after an extended recovery period [62]. These results suggest that atrophy in the precuneus may be more vulnerable to illness severity and more challenging to recover from. Furthermore, a longitudinal study on patients with a short illness duration (<3 years) found that precuneus volume increased and normalized after a short average recovery period of 67 days [11]. This finding further supports the idea that precuneus atrophy is associated with the severity of AN. These findings provide evidence to encourage future research to investigate not only the precuneus but also other brain regions regarding their relationship with changes in disorder severity in AN.

The precuneus has been considered involved in self-related mental representation and may be linked to self-body image evaluation. An fMRI study found that healthy controls showed increased precuneus activation when viewing distorted, fatter self-body images in a body estimation task. However, this modulation was absent in AN [86]. Similarly, another study found that control subjects exhibited greater precuneus activation when viewing self-images than non-self-images, whereas this hyperactivation was absent in AN [87]. This lack of precuneus activation in AN patient during body image processing tasks may indicate impaired estimation of body size, aligning with their tendency to overestimate their body size. The abnormalities found in the precuneus may contribute to body size misperception in AN, reinforcing the drive for weight loss.

## Data Availability

All data produced in the present study are available upon reasonable request to the authors

## VI. Limitations

This meta-analysis has encountered several limitations that were difficult to control. First, most studies recruited adolescents and young adults patients, with scarce data collected from middle-aged or older individuals with AN. Given the potential protective effect age has on GM atrophy in AN, future research should expand the age range to better assess long-term structural outcomes and recovery trajectories in aging populations. Second, sample size in these studies varied and patient recovery status differed, both may introduce variability in reported GM volume changes. Third, studies employed varying neuroimaging techniques (e.g., voxel-based morphometry, cortical thickness analysis), leading to potential discrepancies in GM volume estimation. Lastly, the extent of GM volume normalization over time remains unclear, highlighting the need for longitudinal studies with longer follow-up periods. Future research should consider integrating multimodal neuroimaging approaches to disentangle these influences.

## VI. FUNDING AND ACKNOWLEDGEMENT

This research was funded by the National Institute of Mental Health (NIMH) R01MH106781

## V. Conclusion

This meta-analysis provides a comprehensive examination of GM volume alteration in individuals with AN. Our findings reveal that GM volume loss is evident both in younger and older AN compared to controls, and display a greater magnitude of loss in the younger AN. Older age may be a protective factor against the GM volume loss even after accounting for the effect of AN recovery status. Regional analyses further reveal consistent GM atrophy in key brain structures, including the hippocampus, amygdala, and precuneus, which are critical for cognitive control, emotional regulation, and body image perception. The hippocampus and amygdala also displayed structural changes that correlated with AN symptom, illness duration, and recovery, reinforcing the importance of early intervention to prevent these neurobiological deficits. Our study underscores the vulnerability of GM changes in individuals with AN, and reveals older age as a potential protective factor for GM atrophy. Identification of protective dietary and behavioral factors for brain aging may lead to improved outcomes and successful aging for patients with AN.

